# Occupational stressors, perceived injustice, and pathways to pain interference and insomnia among workers with headache or low back pain

**DOI:** 10.64898/2026.09.18.26363293

**Authors:** Keiko Yamada, Akira Mibu, Tomonori Adachi, Kiyoka Enomoto, Tomohiko Nishigami, Michael Sullivan

## Abstract

**Objective:** Perceived injustice is a psychosocial risk factor associated with delayed recovery and poor occupational outcomes among workers with pain conditions. We examined a theory-informed pathway linking occupational stressors, perceived injustice, pain interference, depressive symptoms, and insomnia, while evaluating selected measurement properties of the Japanese Injustice Experience Questionnaire–Short Form (IEQ-SF-J).

**Methods:** This cross-sectional online survey included 600 Japanese workers (300 with headache and 300 with low back pain). Structural validity of the IEQ-SF-J was evaluated using exploratory and confirmatory factor analyses (CFA) and item response theory. Construct validity was examined using Spearman correlations. Structural equation modelling (SEM) estimated indirect associations between job demands or low job control and pain interference, depressive symptoms, and insomnia, with perceived injustice specified as a latent intervening construct.

**Results:** CFA supported a unidimensional structure (comparative fit index = 0.993; root mean square error of approximation = 0.080). Internal consistency was acceptable (Cronbach’s α = 0.82; McDonald’s ω = 0.90), and test–retest reliability was moderate (intraclass correlation coefficient = 0.60). IEQ-SF-J scores correlated significantly with pain interference, insomnia, and depressive symptoms. Job demands and low job control showed significant indirect associations with all three outcomes involving perceived injustice.

**Conclusions:** The IEQ-SF-J showed supportive evidence for several measurement properties and occupied a theoretically coherent position in associations linking occupational stressors with pain interference, depressive symptoms, and insomnia among workers with headache or low back pain.

## 1. Introduction

Chronic pain among working-age adults is associated with reduced work performance, absenteeism, and longer-term disability, and thus represents a significant occupational and public health concern. Musculoskeletal symptoms are among the leading causes of presenteeism in Japan, imposing a substantial economic burden on the workforce ^1^. In addition, pain severity has been linked to both absenteeism and presenteeism among Japanese full-time workers ^2^. Research on occupational low back pain in Japan has also identified psychosocial factors as important correlates of pain that interfere with work ^3^.

Perceived injustice has emerged as a key psychosocial factor that is particularly relevant to pain-related disability. Sullivan et al. defined perceived injustice as a cognitive appraisal characterized by a sense of severe and irreparable loss and the attribution of blame to others for one’s suffering ^4^. Originally studied in traumatic and occupational injury contexts, perceived injustice has since been associated with poorer recovery and pain-related outcomes in conditions including whiplash injury, work-related low back pain, fibromyalgia, and other persistent musculoskeletal disorders ^4–7^. Notably, these associations remain significant even after accounting for other psychosocial risk factors such as catastrophizing, fear of pain, and depression ^8^.

Sullivan et al. developed the 12-item Injustice Experience Questionnaire (IEQ), which has demonstrated supportive measurement properties in chronic pain and disability populations^4^. To improve feasibility for screening and repeated measurement, the authors later created the 5-item Injustice Experience Questionnaire–Short Form (IEQ-SF), which has a simplified 3-point response scale. This instrument demonstrated acceptable internal consistency in work-disabled individuals with musculoskeletal conditions and major depressive disorder, correlated meaningfully with pain, depressive symptoms, and disability, and exhibited responsiveness to treatment-related change^9^.These findings support the use of the IEQ-SF as an efficient, practical measure of injustice appraisals across disabling health conditions.

While the original 12-item IEQ has been translated into Japanese and evaluated for its measurement properties^10^, that earlier evaluation did not incorporate item response theory (IRT), leaving important item-level measurement characteristics, such as item discrimination and threshold parameters, uncharacterized. Moreover, the measurement properties of the Japanese short form have not been evaluated in occupational pain research. The present study therefore aimed to (i) evaluate selected measurement properties of the Japanese IEQ-SF (IEQ-SF-J) in workers with headache or low back pain, and (ii) to examine a theory-informed pathway model linking occupational stressors (job demands and low job control), perceived injustice, pain interference, depressive symptoms, and insomnia (Fig. 2). By extending the measurement-property evaluation of the IEQ-SF-J to include IRT-based item parameter estimation, this study may provide a stronger foundation for biopsychosocial pain research and occupational health studies in Japan.

**Figure 1.**
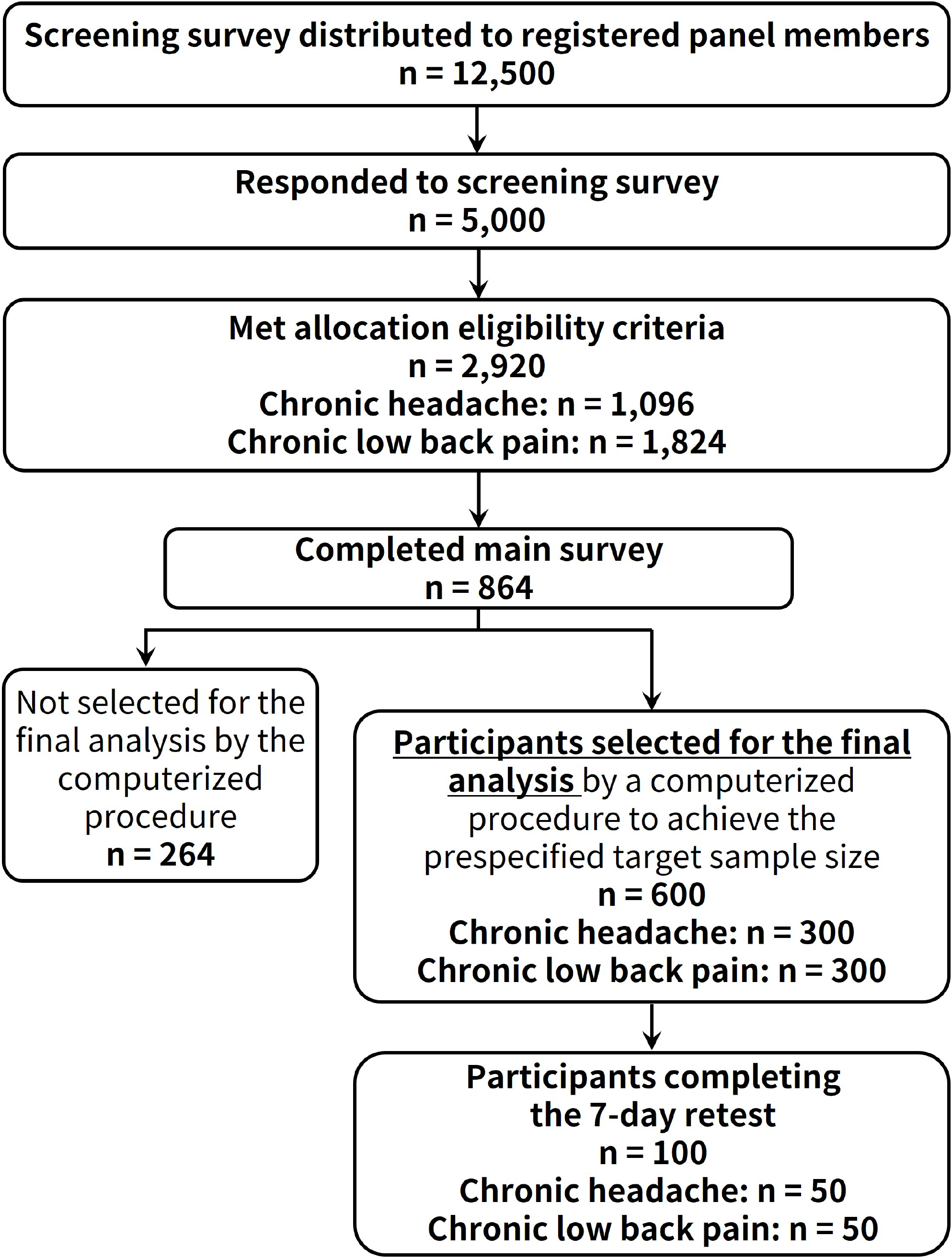
Participant flow diagram. The screening survey was distributed to 12,500 registered panel members, of whom 5,000 responded. Among screening respondents, 2,920 met the allocation eligibility criteria, including 1,096 participants with chronic headache and 1,824 participants with chronic low back pain, irrespective of subsequent completion of the main survey. A total of 864 participants completed the main survey. Because the investigators had prespecified a target sample of 600 participants based on the study design and available budget, the survey provider used a computerized procedure to select 600 participants for the final analysis, comprising 300 participants with chronic headache and 300 with chronic low back pain. The remaining 264 main-survey completers were not selected because the prespecified target sample size had been reached. The retest survey was completed 7 days later by 100 participants (50 with chronic headache and 50 with chronic low back pain). Detailed reason-specific counts for non-completion between screening eligibility and main-survey completion were unavailable from the panel provider.

**Figure 2.**
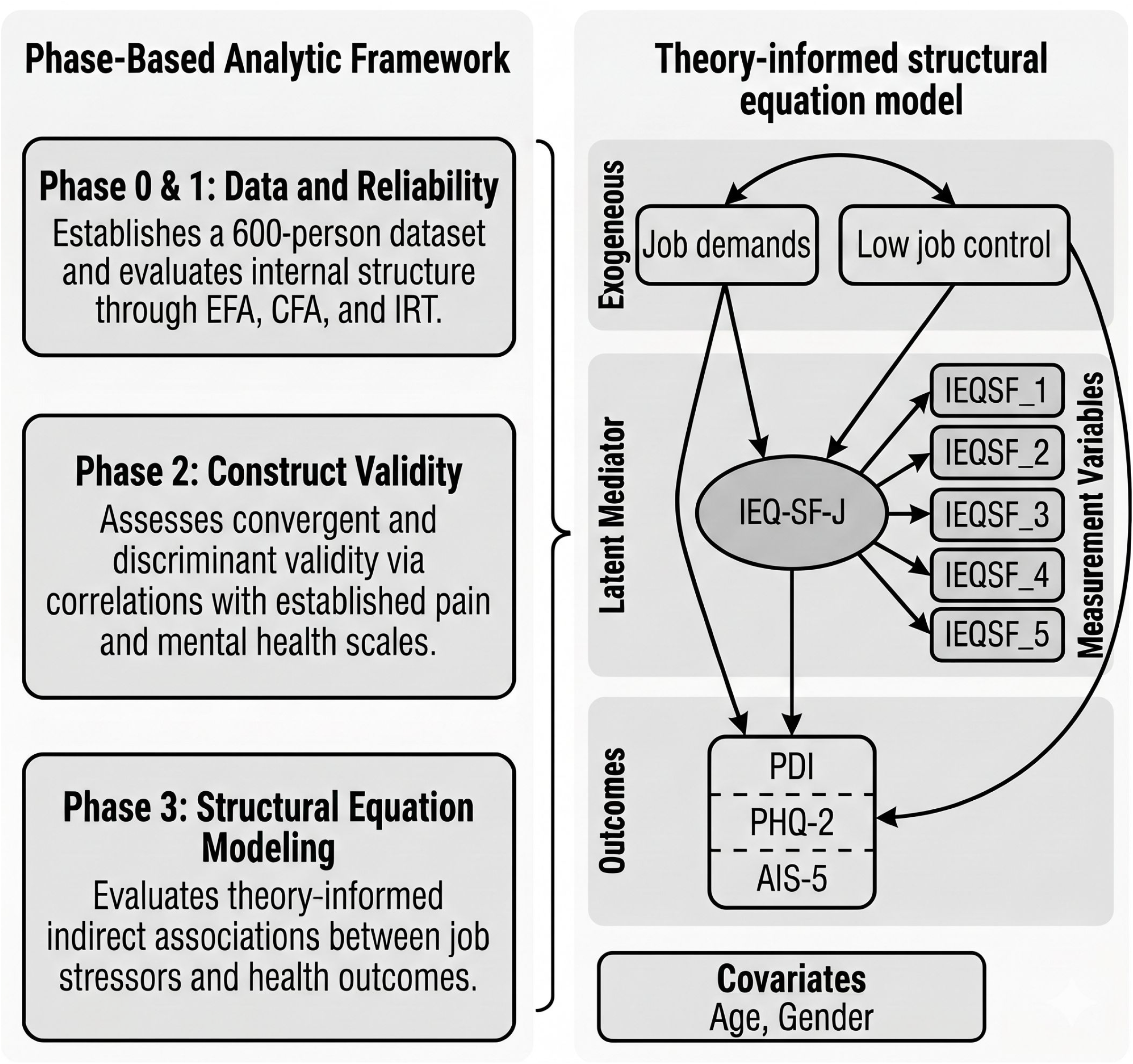
Phase-based analytic framework and theory-informed structural equation model. The left panel summarizes the three analytic phases: Phase 0 & 1, data establishment and evaluation of internal structure using EFA, CFA, and IRT; Phase 2, assessment of construct validity; and Phase 3, evaluation of theory-informed indirect associations between occupational stressors and health outcomes. The right panel shows the structural equation model. Job demands and low job control were specified as exogenous predictors; perceived injustice was modeled as a latent construct indicated by the five IEQ-SF-J items; PDI, PHQ-2, and AIS-5 were specified as outcomes; and age and gender were included as covariates. Abbreviations: AIS-5, Athens Insomnia Scale 5-item version; CFA, confirmatory factor analysis; EFA, exploratory factor analysis; IEQ-SF-J, Japanese version of the shortform Injustice Experience Questionnaire; IRT, item response theory; PDI, Pain Disability Index; PHQ-2, Patient Health Questionnaire-2; SEM, structural equation modeling.

## 2. Methods

### 2.1 Study Design and Participants

This cross-sectional study used data from the 2020 Japanese Biopsychosocial Assessment of Pain project (DC-JBAP2020), a web-based survey previously used to validate the Japanese Pain Disability Index ^11^. Participants were recruited through a commercial internet research panel and were limited to currently employed Japanese adults (aged 20–64) experiencing chronic pain. Consistent with the DC-JBAP2020 report, chronic low back pain was defined as pain persisting for at least 3 months, and chronic daily headache as headache occurring ≥ 15 days per month for ≥ 3 months ^11^. The screening survey was distributed to 12,500 registered panel members, of whom 5,000 responded. Among screening respondents, 2,920 met the allocation eligibility criteria, including 1,096 participants with chronic headache and 1,824 with chronic low back pain, irrespective of subsequent completion of the main survey. A total of 864 participants completed the main survey. Because the investigators had prespecified a target sample of 600 participants based on the study design and available budget, the survey provider used a computerized procedure to select 600 participants for the final analysis, comprising 300 with chronic headache and 300 with chronic low back pain. The remaining 264 main-survey completers were not selected because the prespecified target sample size had been reached. Detailed reason-specific counts for non-completion between screening eligibility and main-survey completion were unavailable from the panel provider (Fig. 1). To evaluate test–retest reliability, a subsample of 100 participants (50 with headache and 50 with low back pain) completed a follow-up IEQ-SF-J 7 days after completing the initial survey ^11^. The sample size was determined by the design of the DC-JBAP2020 survey, which recruited equal-sized headache and low back pain subgroups for evaluation of measurement properties and subgroup comparison. No formal a priori power calculation was performed. The study protocol was approved by the Research Ethics Committee, Faculty of Medicine, Juntendo University (approval number 2020175), and all participants provided web-based informed consent.

### 2.2 Measures

*IEQ-SF-J*. The IEQ-SF is a 5-item short form of the original 12-item IEQ, which was designed to assess perceived injustice in the context of pain and disability ^4,9^. For the present study, we used the linguistically validated IEQ-SF-J that had been produced through forward-back translation, expert review, and cognitive debriefing, strictly adhering to ISPOR good-practice guidelines ^12,13^. Items are rated on a 3-point scale from 0 (“never”) to 2 (“often”), resulting in a total score between 0 and 10. The Japanese item wording and response options are presented in Supplementary Table S2. Copyright © 2002 Michael JL Sullivan, who is also a co-author of the present study. The IEQ-SF is licensed and distributed by Mapi Research Trust on behalf of the copyright holder, and the IEQ-SF-J was developed under license from the copyright holder. Permissions and conditions of use are available through ePROVIDE (https://eprovide.mapi-trust.org). The IEQ-SF-J was specified as the latent construct in the theory-informed pathway model shown in Fig. 2.

#### IEQ

To support hypothesis testing for construct validity, the full 12-item IEQ was administered as a comparator instrument^4^. The Japanese version has previously been evaluated for its measurement properties^10^.

#### Pain Catastrophizing Scale (PCS)

Catastrophic thinking about pain was assessed using the Japanese version of 13-item PCS ^14,15^, which yields a total score ranging from 0 to 52.

*Numeric Rating Scale (NRS)*. Current pain intensity was measured on an 11-point NRS (0–10), a widely used pain assessment scale ^16^.

#### Pain Disability Index (PDI)

Pain-related interference was assessed using the PDI ^17^. We employed the 5-item Japanese scoring format (yielding a total score of 0–50) derived from prior validation work using the DC-JBAP2020 dataset ^11^.

*Athens Insomnia Scale, 5-item version (AIS-5)*. Insomnia symptoms during the preceding month were assessed using a 5-item version of the Athens Insomnia Scale, originally based on ICD-10 criteria ^18^. The Japanese AIS has previously been validated ^19^, and we used a score of ≥ 4 as the risk threshold for clinically relevant insomnia, consistent with cut-off values established in Japanese chronic pain patients ^19,20^.

#### Patient Health Questionnaire-2 (PHQ-2)

Depressive symptoms were assessed using the PHQ-2, a 2-item measure of depressed mood and anhedonia ^21^. A score of ≥ 2 was used as the risk threshold for clinically relevant depressive symptoms, in accordance with prior validation work ^21^.

*Job demands and job control*. Occupational stressors were assessed utilizing 3-item subscales that capture psychological job demands and job control, derived from the Job Content Questionnaire developed by Karasek et al. ^22^. For the pathway analyses, job control was reverse-scored, with higher scores indicating lower perceived job control.

### 2.3 Statistical analysis

The evaluation of measurement properties was conducted in four sequential phases. First, item-level descriptive statistics were examined. Second, dimensionality was evaluated via exploratory factor analysis (EFA), with factor retention determined by visual inspection of the scree plot alongside parallel analysis. This was followed by confirmatory factor analysis (CFA). Given the ordinal nature of the IEQ-SF-J items, the CFA used the weighted least squares mean and variance adjusted (WLSMV) estimator. Model fit was assessed using the comparative fit index (CFI), Tucker–Lewis index (TLI), root mean square error of approximation (RMSEA), and standardized root mean square residual (SRMR), in accordance with established structural equation modelling (SEM) recommendations ^23,24^. Item response theory (IRT) analyses were then conducted using a graded response model to estimate discrimination and threshold parameters. Internal consistency was evaluated using both Cronbach’s α and McDonald’s categorical omega (ω). Categorical omega was calculated based on the standardized factor loadings and residual variances from the WLSMV-based CFA, as it provides a more robust and accurate estimate of reliability for ordinal data by not relying on the assumption of tau-equivalence. The intraclass correlation coefficient (ICC[3,1]) was used to determine test–retest reliability ^25^. Construct validity was evaluated using Spearman rank correlations (with 95% confidence intervals) between IEQ-SF-J and IEQ, PCS, PHQ-2, NRS, PDI, and AIS-5 scores. Finally, a theory-informed SEM estimated direct, indirect, and total associations between job stressors (job demands and low job control) and pain interference, insomnia, and depressive symptoms, with perceived injustice modeled as a latent intervening construct indicated by the five IEQ-SF-J items. All models were adjusted for age and gender. Direct, indirect, and total associations were estimated using bias-corrected bootstrap resampling with 5,000 draws to obtain 95% confidence intervals. All analyses were performed on the total sample and by pain subgroup (headache and low back pain), with statistical significance set at α = 0.05 (two-tailed).

Because no direct global rating of change was collected at retest, sensitivity analyses of test–retest reliability were conducted in participants with stable pain intensity (absolute NRS change ≤1 point) and, more stringently, in participants who additionally showed stable pain-related disability (absolute change ≤1 point in the PDI-5 mean item score).

Measurement invariance across the headache and low back pain subgroups was additionally assessed at the item level using ordinal logistic regression differential item functioning (DIF) analyses. For each IEQ-SF-J item, pain subgroup and its interaction with the four-item rest score were sequentially added to assess uniform and non-uniform DIF. Overall DIF p values were adjusted using the Benjamini–Hochberg false discovery rate procedure, and DIF magnitude was quantified using change in Nagelkerke’s R^2^.

## 3. Results

### 3.1 Participant Characteristics

Table 1 presents the participants’ key characteristics, with detailed sociodemographic and occupational characteristics provided in Supplementary Table S1. The total sample (N = 600) had a mean age of 49.1 years (SD 8.9), with women constituting 36.5% of the cohort. Notably, the headache group had a higher proportion of women (48.7%) than the low back pain group (24.3%). University-level education was the most common educational attainment (42.5%), and desk work accounted for 52.0% of employment in both subgroups, with 63.7% of participants working ≥40 hours per week. Insomnia risk (AIS-5 ≥ 4) was highly prevalent, affecting 75.7% of the total sample; this was more pronounced among those in the headache group (83.7%) than among those with low back pain (67.7%). Risk for depressive symptoms (PHQ-2 ≥ 2) was identified in 27.2% of the overall sample (headache: 37.3%; low back pain: 17.0%). Mean IEQ-SF-J scores were 4.1 (SD 2.5) in the headache group and 3.1 (SD 2.2) in the low back pain group, suggesting a somewhat higher perceived injustice burden among individuals with chronic headache.

**Table 1.** Participant characteristics and key study variables.

|  | **Total (N = 600)** | **Headache (n = 300)** | **Low back pain (n = 300)** |
| --- | --- | --- | --- |
| **Age, years, mean (SD)** | 49.1 (8.9) | 46.7 (8.9) | 51.4 (8.2) |
| **Women, n (%)** | 219 (36.5) | 146 (48.7) | 73 (24.3) |
| **Desk work (office/computer), n (%)** | 312 (52.0) | 156 (52.0) | 156 (52.0) |
| **Weekly working hours $\geq 40$ h/week, n (%)** | 382 (63.7) | 180 (60.0) | 202 (67.3) |
| **Job demands, mean (SD): 3–12** | 8.1 (2.2) | 8.4 (2.2) | 7.8 (2.2) |
| **Job control, mean (SD): 3–12** | 8.0 (2.4) | 7.8 (2.3) | 8.3 (2.3) |
| **IEQ-SF-J, mean (SD): 0–10** | 3.6 (2.4) | 4.1 (2.5) | 3.1 (2.2) |
| **Pain intensity, NRS, mean (SD): 0–10** | 4.8 (1.9) | 4.9 (2.0) | 4.7 (1.9) |
| **Pain interference, PDI, mean (SD): 0–50** | 23.0 (14.9) | 26.2 (15.8) | 19.9 (13.2) |
| **AIS-5, mean (SD):** | 5.7 (3.1) | 6.5 (3.2) | 4.9 (2.8) |
| **Insomnia risk, AIS-5 $\geq 4$, n (%)** | 454 (75.7) | 251 (83.7) | 203 (67.7) |
| **PHQ-2, mean (SD): 0–2** | 0.7 (0.9) | 1.0 (0.9) | 0.5 (0.8) |
| **Depressive symptom risk, PHQ-2 $\geq 2$, n (%)** | 163 (27.2) | 112 (37.3) | 51 (17.0) |

**Abbreviation.** AIS-5, Athens Insomnia Scale 5-item version; IEQ-SF-J, Japanese short-form Injustice Experience Questionnaire; NRS, Numerical Rating Scale; PDI, Pain Disability Index; PHQ-2, Patient Health Questionnaire-2; SD, standard deviation.
**Note.** Continuous variables are presented as mean (SD), and categorical variables as n (%). Weekly working hours $\geq 40$ h/week was calculated by combining the 40–48, 49–59, 60–69, and $\geq 70$ h/week categories. Detailed sociodemographic and occupational characteristics are provided in Supplementary Table S1.

### 3.2 CFA Reliability and Fit Indices

The CFA results are presented in Supplementary Table S2. In the total sample, the unidimensional model demonstrated acceptable fit: CFI = 0.993, TLI = 0.986, RMSEA = 0.080 (90% CI: 0.049–0.113), and SRMR = 0.025. Model fit was excellent in the headache subgroup (CFI = 0.998, TLI = 0.996, RMSEA = 0.043 [90% CI: 0.000–0.098], SRMR = 0.019) and adequate in the low back pain subgroup (CFI = 0.987, TLI = 0.974, RMSEA = 0.103 [90% CI: 0.060–0.151], SRMR = 0.038). Standardized factor loadings were uniformly large and statistically significant across all items and groups, ranging from 0.682 (item 3, headache) to 0.934 (item 5, headache). Internal consistency was excellent when assessed with McDonald’s categorical omega (ω = 0.895 for the total sample; 0.892 for headache; 0.894 for low back pain) and acceptable according to conventional Cronbach’s α (α = 0.82 for the total sample; 0.83 for headache; 0.81 for low back pain). Test–retest reliability over the approximately 7-day interval yielded an ICC (3,1) of 0.60 (95% CI: 0.46–0.71) overall, with pain subgroup values of 0.55 (headache) and 0.64 (low back pain).

Sensitivity analyses restricted to participants with stable pain intensity, and additionally stable pain-related disability, are presented in Supplementary Table S3.

### 3.3 Construct Validity

Supplementary Table S4 presents the Spearman correlations between IEQ-SF-J scores and the external measures. In both pain subgroups, the IEQ-SF-J showed strong correlations with the full IEQ (headache: r = 0.60 [95% CI: 0.52–0.66]; low back pain: r = 0.67 [95% CI: 0.60–0.73]) and the PCS (headache: r = 0.63 [95% CI: 0.55–0.69]; low back pain: r = 0.71 [95% CI: 0.65–0.77]). Moderate positive correlations were observed with the PDI (headache: r = 0.45 [95% CI: 0.35–0.53]; low back pain: r = 0.55 [95% CI: 0.46–0.62]) and the AIS-5 (headache: r = 0.36 [95% CI: 0.25–0.45]; low back pain: r = 0.31 [95% CI: 0.20–0.41]). Aligning with the anticipated discriminant pattern, correlations with job demands and low job control were positive but weaker. Significant, moderate correlations with the PHQ-2 indicate that perceived injustice is meaningfully related to, but conceptually distinct from, depressive symptoms.

### 3.4 Item-Level Descriptives, IRT, and Measurement Invariance

Item-level statistics and IRT parameters are presented in Supplementary Table S5. In the total sample, item means ranged from 0.51 (item 5: “Nothing will ever make up for all that I have gone through”) to 1.05 (item 1: “Most people don’t understand how severe my condition is”). Floor effects (≥15% selecting “never”) were observed for most items, particularly for items 2–5, consistent with the anticipated low endorsement of extreme injustice appraisals within a community-based working population. A ceiling effect was noted only for item 1 in the headache subgroup. EFA factor loadings ranged from 0.622 (Item 1) to 0.818 (Item 5).

Prior to conducting confirmatory analyses, we evaluated the number of factors to retain. Visual inspection of the scree plot revealed a clear, steep decline in eigenvalues after the first factor across the total sample and both pain subgroups (Fig. S1). This unidimensional structure was strongly supported by the parallel analysis (Fig. S2). In all three groups, only the first observed eigenvalue (ranging from 2.80 to 2.95) substantially exceeded its corresponding simulated critical value (1.15).

Conversely, the observed eigenvalues for the second factor (ranging from 0.65 to 0.80) fell well below the simulated threshold (1.10). These convergent results supported the retention of a single-factor structure for subsequent CFA and IRT analyses.

The IRT graded response model parameters demonstrated high discrimination across all items, with discrimination values (a) ranging from 1.82 (item 3) to 4.62 (item 5) in the total sample. Item 5 was particularly informative; its exceptionally high discrimination parameter (a = 4.62) indicates excellent ability to differentiate individuals along the latent continuum of perceived injustice. Threshold parameters (b_1_) ranged from −1.44 for item 1, suggesting it is relatively easy to endorse at least “sometimes”, to 0.19 for item 5, indicating it requires a higher level of the latent trait to endorse. The b_2_ values ranged from 1.17 to 1.88, appropriately positioning the “often” threshold in the upper portion of the latent continuum.

Item-level DIF analyses identified statistically detectable DIF after false-discovery-rate correction for Items 1–3 (q = 0.0102, <0.0001, and 0.0102, respectively), whereas Items 4 and 5 showed no statistical DIF. However, overall ΔNagelkerke R^2^ values were small for all five items (range, 0.0010–0.0279; maximum for Item 2), indicating limited practical impact and supporting approximate item-level invariance across the two pain subgroups (Supplementary Table S5).

### 3.5 Theory-Informed Pathway Model: SEM Results

The main indirect associations in the overall sample are summarized in Table 2, and the full SEM results, including direct effects, total effects, model fit indices, and subgroup analyses, are provided in Supplementary Table S6. Model fit was acceptable across the specified models (RMSEA range: 0.043–0.103; CFI range: 0.974–0.991).

**Table 2.** Main indirect associations between occupational stressors and outcomes involving perceived injustice in the overall sample.

| Outcome | Job demands: indirect association involving perceived injustice | Low job control: indirect association involving perceived injustice | Summary interpretation |
| --- | --- | --- | --- |
| Pain interference | $\beta = 0.133$, 95% CI 0.073 to 0.192 | $\beta = 0.082$, 95% CI 0.028 to 0.142 | Both job demands and low job control showed indirect associations with pain interference involving perceived injustice. |
| Insomnia | $\beta = 0.089$, 95% CI 0.049 to 0.135 | $\beta = 0.055$, 95% CI 0.018 to 0.100 | Both job demands and low job control showed indirect associations with insomnia involving perceived injustice. |
| Depressive symptoms | $\beta = 0.084$, 95% CI 0.046 to 0.130 | $\beta = 0.052$, 95% CI 0.017 to 0.092 | Both occupational stressors showed indirect associations with depressive symptoms involving perceived injustice. |

**Abbreviations:** CI, confidence interval; IEQ-SF-J, Japanese version of the short-form Injustice Experience Questionnaire.
**Note:** Indirect associations are presented as standardized coefficients with 95% bias-corrected bootstrap confidence intervals based on 5,000 resamples. Higher low job control scores indicate lower perceived job control. In the structural equation models, perceived injustice was specified as a latent construct indicated by the five IEQ-SF-J items. Full direct effects, total effects, model fit indices, and subgroup analyses are provided in Supplementary Table S6.

In the overall sample, both job demands and low job control showed significant indirect associations with pain interference involving perceived injustice. The standardized indirect association was β = 0.133 (95% bias-corrected bootstrap CI: 0.073 to 0.192) for job demands and β = 0.082 (95% CI: 0.028 to 0.142) for low job control. Similar patterns were observed for insomnia, with significant indirect associations for job demands (β = 0.089, 95% CI: 0.049 to 0.135) and low job control (β = 0.055, 95% CI: 0.018 to 0.100). For depressive symptoms, job demands (β = 0.084, 95% CI: 0.046 to 0.130) and low job control (β = 0.052, 95% CI: 0.017 to 0.092) also showed significant indirect associations involving perceived injustice. Subgroup analyses are presented in Supplementary Table S6; these results suggested broadly similar directions of association, although several subgroup-specific estimates should be interpreted cautiously.

## 4. Discussion

To our knowledge, this study provides the first evaluation of selected measurement properties of the IEQ-SF-J in Japanese workers reporting headache or low back pain during the preceding 4 weeks. Across sequential analytic phases, our findings provide supportive evidence for several measurement properties of the IEQ-SF-J, a brief instrument for measuring perceived injustice. Multiple extraction criteria, including scree plot trajectories and parallel analysis, consistently support a unidimensional structure. Furthermore, the pathway analyses suggest that perceived injustice occupies a theoretically coherent position in the associations between occupational stressors, pain interference, depressive symptoms, and insomnia. These findings extend prior work on perceived injustice in pain populations, which has consistently linked injustice appraisals to pain-related interference and adverse pain-related outcomes ^4,6,26^.

### 4.1 Measurement properties of the IEQ-SF-J

The IEQ-SF-J’s unidimensional structure was supported across both headache and low back pain groups, aligning with the original conceptualization of perceived injustice as a unitary appraisal construct in the IEQ literature ^4^. The internal consistency estimates observed in this study, particularly the robust categorical omega values (ω > 0.89), indicate excellent internal consistency that accounts for the ordinal nature of the 3-point response scale. These results closely mirror the original IEQ-SF measurement-property results ^9^, which confirmed acceptable reliability among work-disabled individuals with musculoskeletal conditions and major depressive disorder. These findings indicate that favorable measurement characteristics of the short form are also observed in the Japanese version and within a working population that likely represents less severe and more heterogeneous pain states than those typically found in clinical rehabilitation settings.

While the CFA fit was generally acceptable, the RMSEA was somewhat elevated in a segment of the sample. This should be interpreted with caution within the context of a very short, single-factor model; RMSEA can overreject close-fitting models with limited degrees of freedom and should not be relied upon as an isolated, universal threshold ^24^. The more favorable CFI and SRMR values support the conclusion that the scale is adequately represented by a one-factor solution.

The IRT findings were likewise informative. Item 5, referencing the irreparability of loss demonstrated the highest discrimination, corroborating earlier work that identifies severe and irreversible loss as a core dimension of injustice appraisal ^4^. Conversely, the item regarding the failure of others to understand the severity of one’s condition functioned at a lower threshold. This indicates the item’s sensitivity to the comparatively mild injustice appraisals typical of community samples. The floor tendency of several items was anticipated, given that the sample comprised actively employed adults rather than clinical rehabilitation patients, among whom perceived injustice is generally more pronounced ^4,6^.

Test–retest reliability over the approximately 7-day interval was moderate, falling below the coefficient reported in the original IEQ-SF study ^9^. This discrepancy can likely be attributed to several factors, including the present study’s use of an online survey format, its broader population-based case mix, and the relatively small retest subsample. Interpreted in accordance with current reporting conventions, the ICC denotes moderate reliability rather than robust temporal stability ^25^. In practice, these results suggest that the IEQ-SF-J is well-suited for cross-sectional epidemiologic research, whereas its utility for short-interval longitudinal monitoring may require further empirical scrutiny.

### 4.2 Construct validity

Construct validity was supported by the observed correlation matrix. The strong association with the full IEQ provides convergent evidence supporting construct validity, confirming that the IEQ-SF-J successfully captures the core content of the original instrument ^4^. Its moderate association with pain catastrophizing is consistent with prior research establishing that perceived injustice and catastrophizing are related yet distinct psychosocial constructs in the context of pain ^6,26^. Furthermore, correlations with pain intensity, pain interference, insomnia, and depressive symptoms underscore the IEQ-SF-J’s clinical relevance while confirming that it is not merely a proxy for generalized emotional distress.

The association with pain interference is particularly noteworthy. Using the same DC-JBAP2020 platform previously employed for the Japanese validation of the PDI—but with broadened eligibility criteria that included workers with recent pain irrespective of chronicity—our finding of convergence between the IEQ-SF-J and the PDI aligns with the broader injustice literature and prior Japanese validation work on pain-related interference ^11,17^.

### 4.3 Theory-informed pathway model

A central contribution of this study is the integration of the IEQ-SF-J into a theory-informed occupational pathway model, extending beyond conventional evaluation of measurement properties. Our results suggest that perceived injustice may represent a psychological process through which adverse working conditions are statistically linked to pain interference, depressive symptoms, and insomnia. This interpretation aligns with existing evidence showing that perceived injustice is associated with greater pain-related interference, emotional distress, and maladaptive adjustment across various pain conditions, even when accounting for other psychological risk factors ^4,6,26^. However, given the study’s cross-sectional design, these pathways should be interpreted as theoretically ordered associations rather than definitive causal trajectories.

The observed indirect associations involving pain interference are clinically plausible. Occupational stressors, such as high demands or low control, may be associated with stronger perceptions that pain-related losses are unfair, unrecognized, or irreversible. These appraisals may, in turn, be related to greater day-to-day functional interference. This conceptualization is consistent with the original IEQ framework, in which blame, a sense of unfairness, and the irreparability of loss are central cognitive themes associated with pain-related interference ^4^.

The insomnia findings similarly warrant attention. Low job control showed both direct and indirect associations with insomnia in the overall sample, suggesting that perceived injustice may be one of several psychological processes involved in the association between job stressors and sleep disturbance. This finding is consistent with literature showing that headache and insomnia are closely intertwined and frequently co-occur with affective symptoms ^27–29^. In this context, injustice appraisals may be relevant to sleep difficulties in some workers, whereas residual direct associations may reflect other processes, such as physiological arousal, pain burden, or mood-related factors.

The findings for depressive symptoms further suggest that perceived injustice may be relevant not only to pain interference and insomnia but also to affective burden among workers with pain. In the overall sample, both job demands and low job control showed significant indirect associations with depressive symptoms involving perceived injustice. This pattern suggests that depressive symptoms may be related not only to occupational stressors themselves but also to the appraisal of being unfairly burdened despite pain. Although subgroup differences should be interpreted cautiously, they may suggest that the role of low job control differs between workers with headache and those with low back pain.

Longitudinal evidence also indicates that pain and these outcomes can precede one another in directions not represented in our model. Among people with persistent pain, new-onset sleep problems predicted subsequent probable depression, with pain interference contributing modestly to this pathway ^30^. Chronic low back pain prospectively predicted later depressive symptoms in the 1958 British birth cohort ^31^. For headache disorders, migraine predicted later major depressive episodes, while depression also predicted incident migraine in population-based cohorts ^32,33^. Prospective HUNT studies further found headache to predict incident insomnia 11 years later and insomnia to predict later headache, supporting a bidirectional relationship between headache and sleep disturbance ^34,35^. Accordingly, our cross-sectional SEM should be interpreted as one theory-informed ordering of contemporaneous associations rather than evidence that the specified paths represent a unique temporal or causal sequence.

### 4.4 Subgroup heterogeneity

Meaningful differences were observed between the pain subgroups. The headache subgroup exhibited a greater burden of both insomnia and depressive symptoms, consistent with prior research reporting the strong comorbidity among headache disorders, poor sleep architecture, and affective distress ^27,28^. Conversely, results for the low back pain subgroup were relatively more stable in the pain interference model, suggesting that injustice appraisals may represent a particularly interpretable psychosocial pathway for pain-related functional interference within this specific condition. However, the subgroup SEMs relied on smaller effective sample sizes, rendering some estimates statistically unstable, indicating that these subgroup variations should be interpreted judiciously.

### 4.5 Limitations

Several limitations of this study warrant consideration. First, its cross-sectional design precludes definitive conclusions regarding temporal ordering or causality. Second, participants were recruited via a commercial online panel, which may constrain generalizability and introduce selection bias. Third, the retest subsample was small, thereby limiting the precision of the ICC estimate. Fourth, certain model-fit and bootstrap estimates—particularly in the subgroup analyses—were borderline or statistically unstable and thus require replication in independent cohorts. Fifth, we assessed occupational stressors using brief indicator subscales rather than a comprehensive, multidimensional work-stress battery. Finally, the study’s reliance on self-reported data introduces the potential for common-method variance inflation.

### 4.6 Clinical and research implications

This study’s findings support the use of the IEQ-SF-J as a practical instrument for occupational pain research in Japan. The scale is efficient, easily administered, and now underpinned by initial evidence of structural validity, internal consistency, and construct validity among Japanese workers with recent headache or low back pain. Given the original English-language scale’s responsiveness to treatment-related change ^9^, the IEQ-SF-J holds promise for epidemiologic screening and as a viable outcome measure in occupational rehabilitation and psychosocial intervention trials. More broadly, these findings support the view that perceived injustice is not merely an immutable individual trait but may also reflect how workers interpret and internalize adverse, constrained occupational circumstances. This perspective has practical implications for interventions targeting workload management, enhancing job control, and implementing psychologically informed pain care.

### 5. Conclusions

The IEQ-SF-J showed evidence supporting several measurement properties in Japanese workers with chronic headache and low back pain. Our findings support structural validity, adequate internal consistency, moderate test-retest reliability, and construct validity based on associations with established pain-related measures, and informative IRT item parameters. Additionally, a theory-informed pathway model suggests that perceived injustice, as measured by the IEQ-SF-J latent construct, occupies a theoretically plausible position in indirect associations linking occupational stressors (job demands and low job control) with pain interference, depressive symptoms, and insomnia. These results support the IEQ-SF-J as a practical instrument for biopsychosocial pain research and occupational health studies in Japan, and they provide a basis for its application in future longitudinal and interventional research.

## Data Availability

The data that support the findings of this study are not publicly available owing to ethical restrictions… However, the data are available from the corresponding author upon reasonable request and if granted permission by the Research Ethics Committee.

## Abbreviations

AIS-5: Athens Insomnia Scale; 5-item version
CFA: confirmatory factor analysis
CFI: comparative fit index
CI: confidence interval
DC-JBAP2020: 2020 Japanese Biopsychosocial Assessment of Pain project
DIF: differential item functioning
EFA: exploratory factor analysis
ICC: intraclass correlation coefficient
ICD-10: International Classification of Diseases, 10th Revision
IEQ: Injustice Experience Questionnaire
IEQ-SF: Injustice Experience Questionnaire–Short Form
IEQ-SF-J: Japanese version of the Injustice Experience Questionnaire–Short Form
IRT: item response theory
ISPOR: International Society for Pharmacoeconomics and Outcomes Research
NRS: Numeric Rating Scale
PCS: Pain Catastrophizing Scale
PDI: Pain Disability Index
PDI-5: 5-item Pain Disability Index scoring format
PHQ-2: Patient Health Questionnaire-2
RMSEA: root mean square error of approximation
SEM: structural equation modelling
SRMR: standardized root mean square residual
TLI: Tucker–Lewis index
WLSMV: weighted least squares mean and variance adjusted.

## Acknowledgments

All authors appreciate Sonora Kogo and Dr. Kenta Wakaizumi for supporting this research. We thank CJ Singleton, PhD, from Edanz (https://jp.edanz.com/ac) for editing a draft of this manuscript.

## Statements and Declarations

### Author contributions

Conceptualization: K.Y.; Data curation: K.Y.; Formal analysis: K.Y.; Funding acquisition: K.Y.; Investigation: K.Y.; Methodology: K.Y., A.M., T.A., K.E., T.N., and M.S. (Japanese translation and/or cross-cultural adaptation of the IEQ-SF-J); Software: K.Y.; Visualization: K.Y.; Writing – original draft: K.Y.; Writing – review & editing: K.Y., A.M., T.A., K.E., T.N., and M.S. All authors have read and agreed to the published version of the manuscript.

### Ethics approval

This study adhered to the ethical principles outlined in the Declaration of Helsinki of 1975, as revised in 2013. The study protocol was approved by the Research Ethics Committee, Faculty of Medicine, Juntendo University (approval number 2020175).

### Consent to participate

All participants provided their web-based informed consent before responding to the online questionnaire

### Data availability

The data that support the findings of this study are not publicly available owing to ethical restrictions—they contain information that could compromise the privacy of research participants. In accordance with the Ethical Guidelines for Medical and Biological Research Involving Human Subjects in Japan, the Research Ethics Committee, Faculty of Medicine, Juntendo University has placed restrictions on the dissemination of these data. However, the data are available from the corresponding author (K.Y.) upon reasonable request and if granted permission by the Research Ethics Committee.

## References

1. Yoshimoto T, Oka H, Fujii T, Nagata T, Matsudaira K. The economic burden of lost productivity due to presenteeism caused by health conditions among workers in Japan. J Occup Environ Med. 2020 Oct;62(10):883–8.

2. Tanaka C, Wakaizumi K, Takaoka S, Matsudaira K, Mimura M, Fujisawa D, et al. A cross-sectional study of the impact of pain severity on absenteeism and presenteeism among Japanese full-time workers. Pain Ther. 2022 Dec;11(4):1179–93.

3. Yoshimoto T, Oka H, Katsuhira J, Fujii T, Masuda K, Tanaka S, et al. Prognostic psychosocial factors for disabling low back pain in Japanese hospital workers. PLoS One. 2017 May 22;12(5):e0177908.

4. Sullivan MJL, Adams H, Horan S, Maher D, Boland D, Gross R. The role of perceived injustice in the experience of chronic pain and disability: scale development and validation. J Occup Rehabil. 2008 Sept;18(3):249–61.

5. Sullivan MJL, Adams H, Martel MO, Scott W, Wideman T. Catastrophizing and perceived injustice: risk factors for the transition to chronicity after whiplash injury. Spine (Phila Pa 1976). 2011 Dec 1;36(25 Suppl):S244–9.

6. Rodero B, Luciano JV, Montero-Marín J, Casanueva B, Palacin JC, Gili M, et al. Perceived injustice in fibromyalgia: psychometric characteristics of the Injustice Experience Questionnaire and relationship with pain catastrophising and pain acceptance. J Psychosom Res. 2012 Aug;73(2):86–91.

7. Scott W, Trost Z, Milioto M, Sullivan MJL. Further validation of a measure of injury-related injustice perceptions to identify risk for occupational disability: a prospective study of individuals with whiplash injury. J Occup Rehabil. 2013 Dec;23(4):557–65.

8. Sullivan MJL, Scott W, Trost Z. Perceived injustice: a risk factor for problematic pain outcomes. Clin J Pain. 2012 July;28(6):484–8.

9. Sullivan MJL, Adams H, Yakobov E, Ellis T, Thibault P. Psychometric properties of a brief instrument to assess perceptions of injustice associated with debilitating health and mental health conditions. Psychol Inj Law. 2016 Mar;9(1):48–54.

10. Yamada K, Adachi T, Mibu A, Nishigami T, Motoyama Y, Uematsu H, et al. Injustice Experience Questionnaire, Japanese version: Cross-cultural factor-structure comparison and demographics associated with perceived injustice. PLoS One. 2016 Aug 3;11(8):e0160567.

11. Yamada K, Mibu A, Kogo S, Sullivan M, Nishigami T. Reliability and validity of the Japanese version of Pain Disability Index. PLoS One. 2022 Sept 12;17(9):e0274445.

12. Wild D, Grove A, Martin M, Eremenco S, McElroy S, Verjee-Lorenz A, et al. Principles of Good Practice for the translation and Cultural Adaptation process for patient-reported outcomes (PRO) measures: Report of the ISPOR task force for translation and Cultural Adaptation. Value Health. 2005 Mar;8(2):94–104.

13. Yamada K, Wakaizumi K, Mibu A, Kogo S, Iseki M, Nishigami T. Development of a Japanese version of the Injustice Experience Questionnaire-Short Form: Translation with linguistic validity (in Japanese). Pain Clinic. 2021;42(2):239–44.

14. Sullivan MJL, Bishop SR, Pivik J. The Pain Catastrophizing Scale: Development and validation. Psychol Assess. 1995;7(4):524–32.

15. Hirofumi Matsuoka YS. Assessment of Cognitive Aspect of Pain : Development, Reliability, and Validation of Japanese Version of Pain Catastrophizing Scale (in Japanese). Shinshin Igaku. 2007 Aug 1;47(2):95–102.

16. Hartrick CT, Kovan JP, Shapiro S. The numeric rating scale for clinical pain measurement: a ratio measure? Pain Pract. 2003 Dec;3(4):310–6.

17. Pollard CA. Preliminary validity study of the pain disability index. Percept Mot Skills. 1984 Dec;59(3):974.

18. Soldatos CR, Dikeos DG, Paparrigopoulos TJ. Athens Insomnia Scale: validation of an instrument based on ICD-10 criteria. J Psychosom Res. 2000 June;48(6):555–60.

19. Okajima I, Nakajima S, Kobayashi M, Inoue Y. Development and validation of the Japanese version of the Athens Insomnia Scale. Psychiatry Clin Neurosci. 2013 Sept;67(6):420–5.

20. Enomoto K, Adachi T, Yamada K, Inoue D, Nakanishi M, Nishigami T, et al. Reliability and validity of the Athens Insomnia Scale in chronic pain patients. J Pain Res. 2018 Apr 16;11:793–801.

21. Kroenke K, Spitzer RL, Williams JBW. The Patient Health Questionnaire-2: validity of a two-item depression screener. Med Care. 2003 Nov;41(11):1284–92.

22. Karasek R, Brisson C, Kawakami N, Houtman I, Bongers P, Amick B. The Job Content Questionnaire (JCQ): an instrument for internationally comparative assessments of psychosocial job characteristics. J Occup Health Psychol. 1998 Oct;3(4):322–55.

23. Hu LT, Bentler PM. Cutoff criteria for fit indexes in covariance structure analysis: Conventional criteria versus new alternatives. Struct Equ Modeling. 1999 Jan;6(1):1–55.

24. Chen F, Curran PJ, Bollen KA, Kirby J, Paxton P. An empirical evaluation of the use of fixed cutoff points in RMSEA test statistic in Structural Equation Models. Sociol Methods Res. 2008 Jan 1;36(4):462–94.

25. Koo TK, Li MY. A guideline of selecting and reporting intraclass correlation coefficients for reliability research. J Chiropr Med. 2016 June;15(2):155–63.

26. Scott W, Trost Z, Bernier E, Sullivan MJL. Anger differentially mediates the relationship between perceived injustice and chronic pain outcomes. Pain. 2013 Sept;154(9):1691–8.

27. Rains JC, Poceta JS. Headache and sleep disorders: review and clinical implications for headache management. Headache. 2006 Oct;46(9):1344–63.

28. Ødegård SS, Engstrøm M, Sand T, Stovner LJ, Zwart JA, Hagen K. Associations between sleep disturbance and primary headaches: the third Nord-Trøndelag Health Study. J Headache Pain. 2010 June;11(3):197–206.

29. Sancisi E, Cevoli S, Vignatelli L, Nicodemo M, Pierangeli G, Zanigni S, et al. Increased prevalence of sleep disorders in chronic headache: a case-control study. Headache. 2010 Oct;50(9):1464– 72.

30. Campbell P, Tang N, McBeth J, Lewis M, Main CJ, Croft PR, et al. The role of sleep problems in the development of depression in those with persistent pain: a prospective cohort study. Sleep. 2013 Nov 1;36(11):1693–8.

31. Dickson C, Zhou A, MacIntyre E, Hyppönen E. Do Chronic Low Back Pain and Chronic Widespread Pain differ in their association with Depression Symptoms in the 1958 British Cohort? Pain Med. 2023 June 1;24(6):644–51.

32. Modgill G, Jette N, Wang JL, Becker WJ, Patten SB. A population-based longitudinal community study of major depression and migraine. Headache. 2012 Mar;52(3):422–32.

33. Swanson SA, Zeng Y, Weeks M, Colman I. The contribution of stress to the comorbidity of migraine and major depression: results from a prospective cohort study. BMJ Open. 2013 Mar 9;3(3):e002057.

34. Ødegård SS, Sand T, Engstrøm M, Zwart JA, Hagen K. The impact of headache and chronic musculoskeletal complaints on the risk of insomnia: longitudinal data from the Nord-Trøndelag health study. J Headache Pain. 2013 Mar 12;14(1):24.

35. Odegård SS, Sand T, Engstrøm M, Stovner LJ, Zwart JA, Hagen K. The long-term effect of insomnia on primary headaches: a prospective population-based cohort study (HUNT-2 and HUNT-3). Headache. 2011 Apr;51(4):570–80.

