## Supplemental Tables and Figures for "Occupational stressors, perceived injustice, and pathways to pain interference and insomnia among workers with headache or low back pain"

**Table S1. Participant characteristics by pain subgroup**

| | Total (N = 600) | Headache (n = 300) | Low back pain (n = 300) |
|---|---|---|---|
| **Age, years, mean (SD)** | 49.1 (8.9) | 46.7 (8.9) | 51.4 (8.2) |
| **Gender, women, n (%)** | 219 (36.5) | 146 (48.7) | 73 (24.3) |
| **Educational attainment, n (%)** | | | |
| Junior high school | 25 (4.2) | 16 (5.3) | 9 (3.0) |
| High school | 136 (22.7) | 74 (24.7) | 62 (20.7) |
| Vocational school | 78 (13.0) | 41 (13.7) | 37 (12.3) |
| Junior school | 58 (9.7) | 36 (12.0) | 22 (7.3) |
| Technical college | 6 (1.0) | 2 (0.7) | 4 (1.3) |
| University | 255 (42.5) | 107 (35.7) | 148 (49.3) |
| Graduate school | 41 (6.8) | 23 (7.7) | 19 (6.3) |
| Other | 1 (0.2) | 1 (0.3) | 0 (0) |
| **Marital status, n (%)** | | | |
| Married | 383 (63.8) | 172 (57.3) | 211 (70.3) |
| Single | 137 (22.8) | 83 (27.7) | 54 (18.0) |
| Divorced | 66 (11.0) | 39 (13.0) | 27 (9.0) |
| Widowed | 14 (2.3) | 6 (2.0) | 8 (2.7) |
| **Job category, n (%)** | | | |
| Agriculture, forestry, and fisheries | 3 (0.5) | 2 (0.7) | 1 (0.3) |
| Mining and construction | 27 (4.5) | 16 (5.3) | 11 (3.7) |
| Manufacturing | 94 (15.7) | 37 (12.3) | 57 (19.0) |
| Electricity, gas, heat supply, and water | 10 (1.7) | 3 (1.0) | 7 (2.3) |
| Information and communications | 23 (3.8) | 14 (4.7) | 9 (3.0) |
| Transport and postal services | 19 (3.2) | 6 (2.0) | 13 (4.3) |
| Wholesale and retail trade | 65 (10.8) | 35 (11.7) | 30 (10.0) |
| Finance and insurance | 15 (2.5) | 12 (4.0) | 3 (1.0) |
| Real estate | 14 (2.3) | 9 (3.0) | 5 (1.7) |
| Accommodations, eating and drinking services | 21 (3.5) | 11 (3.7) | 10 (3.3) |
| Medical, health care and welfare | 92 (15.3) | 52 (17.3) | 40 (13.3) |
| Education and learning support | 37 (6.2) | 16 (5.3) | 21 (7.0) |
| Other services | 107 (17.8) | 54 (18.0) | 53 (17.7) |
| Government, except elsewhere classified | 30 (5.0) | 13 (4.3) | 17 (5.7) |
| Other | 43 (7.2) | 20 (6.7) | 23 (7.7) |
| **Job style, n (%)** | | | |

| | | | |
|---|---|---|---|
| Desk work (office/computer) | 312 (52.0) | 156 (52.0) | 156 (52.0) |
| Interpersonal work (sales/service) | 114 (19.0) | 64 (21.3) | 50 (16.7) |
| Physical work (production/care etc.) | 174 (29.0) | 80 (26.7) | 94 (31.3) |
| **Weekly working hours, n (%)** | | | |
| <40 h/week | 218 (36.3) | 120 (40.0) | 98 (32.7) |
| 40–48 h/week | 270 (45.0) | 121 (40.3) | 149 (49.7) |
| 49–59 h/week | 51 (8.5) | 27 (9.0) | 24 (8.0) |
| 60–69 h/week | 31 (5.2) | 14 (4.7) | 17 (5.7) |
| ≥70 h/week | 30 (5.0) | 18 (6.0) | 12 (4.0) |
| **Firm size, n (%)** | | | |
| <50 employees | 262 (43.7) | 127 (42.7) | 135 (45.0) |
| 50–299 employees | 135 (22.5) | 66 (22.0) | 69 (23.0) |
| 300–899 employees | 76 (12.7) | 45 (15.0) | 31 (10.3) |
| ≥1000 employees | 127 (21.2) | 62 (20.7) | 65 (21.7) |
| **Current smoker, n (%)** | 215 (35.8) | 105 (35.0) | 110 (36.7) |
| **Insufficient exercise, n (%)** | 418 (69.7) | 208 (69.3) | 210 (70.0) |
| **IEQ-SF-J, mean (SD): 0–10** | 3.6 (2.4) | 4.1 (2.5) | 3.1 (2.2) |
| **IEQ, mean (SD): 0–48** | 16.8 (12.8) | 19.2 (13.1) | 14.4 (12.0) |
| **PCS, mean (SD): 0–52** | 26.8 (11.2) | 28.6 (11.2) | 25.0 (10.9) |
| **PHQ-2 risk (≥2), n (%)** | 163 (27.2) | 112 (37.3) | 51 (17.0) |
| **PHQ-2, mean (SD): 0–2** | 0.7 (0.9) | 1.0 (0.9) | 0.5 (0.8) |
| **Job demands, mean (SD): 3–12** | 8.1 (2.2) | 8.4 (2.2) | 7.8 (2.2) |
| **Job control, mean (SD): 3–12** | 8.0 (2.4) | 7.8 (2.3) | 8.3 (2.3) |
| **NRS, mean (SD): 0–10** | 4.8 (1.9) | 4.9 (2.0) | 4.7 (1.9) |
| **PDI, mean (SD): 0–50** | 23.0 (14.9) | 26.2 (15.8) | 19.9 (13.2) |
| **AIS-5 insomnia risk (≥4), n (%)** | 454 (75.7) | 251 (83.7) | 203 (67.7) |
| **AIS-5, mean (SD):** | 5.7 (3.1) | 6.5 (3.2) | 4.9 (2.8) |

**Abbreviation.** AIS-5, Athens Insomnia Scale 5-item version; NRS, Numerical Rating Scale; IEQ, Injustice Experience Questionnaire; IEQ-SF-J, Japanese version of the short-form Injustice Experience Questionnaire; PCS, Pain Catastrophizing Scale; PDI, Pain Disability Index; PHQ-2, Patient Health Questionnaire-2.
**Note.** Continuous variables are presented as mean (SD), categorical variables as n (%).

**Table S2. CFA fit indices and factor loadings of IEQ-SF-J by pain subgroup**

| | Total, n = 600 | Headache, n = 300 | Low back pain, n = 300 |
|---|---|---|---|
| **Reliability** | | | |
| McDonald's categorical ω | 0.90 | 0.89 | 0.89 |
| Cronbach's α | 0.82 | 0.83 | 0.81 |
| ICC(3,1) [95% CI] | 0.60 [0.46, 0.71] | 0.55 [0.33, 0.72] | 0.64 [0.44, 0.78] |
| **Sampling Adequacy** | | | |
| KMO measure | 0.83 | 0.84 | 0.81 |
| **Fit Indices** | | | |
| χ2 (df) | 24.021 (5) | 7.753 (5) | 21.062 (5) |
| p-value | < 0.001 | 0.17 | 0.001 |
| CFI | 0.993 | 0.998 | 0.987 |
| TLI | 0.986 | 0.996 | 0.974 |
| RMSEA [90% CI] | 0.080 [0.049, 0.113] | 0.043 [0.000, 0.098] | 0.103 [0.060, 0.151] |
| SRMR | 0.025 | 0.019 | 0.038 |
| **Standardized Factor Loadings (SE)** | | | |
| IEQSF1: 私がどれほどつらいかわかってくれる人はほとんどいない。 | 0.748 (0.025) | 0.774 (0.030) | 0.752 (0.048) |
| IEQSF2: 私の人生はもう元には戻ら | 0.763 (0.024) | 0.773 (0.032) | 0.721 (0.040) |
| IEQSF3: こんな人生には納得がいかない。 | 0.713 (0.027) | 0.682 (0.039) | 0.728 (0.041) |
| IEQSF4: このことが自分の身に起こったと信じられない。 | 0.811 (0.021) | 0.767 (0.032) | 0.856 (0.030) |
| IEQSF5: このひどい経験は他のことで埋め合わせができない。 | 0.923 (0.018) | 0.934 (0.026) | 0.896 (0.027) |

**Abbreviation.** CFI, comparative fit index; CI, confidence interval; ICC, intraclass correlation coefficient; KMO, Kaiser–Meyer–Olkin measure of sampling adequacy; RMSEA, root mean square error of approximation; SE, standard error; SRMR, standardized root mean square residual; TLI, Tucker–Lewis index.

**Note.** Response options were 0 = never, 1 = sometimes, and 2 = often. The corresponding Japanese response options were 0 = まったくそう思わない, 1 = ときどきそう思う, and 2 = よくそう思う. Total scores range from 0 to 10, with higher scores indicating greater perceived injustice. $\chi^2$ chi-square value based on the weighted least squares mean and variance adjusted (WLSMV) estimator. All factor loadings were significant at $p < 0.001$. ICC(3,1) for test-retest reliability was calculated from data collected from 100 participants (headache n = 50, low back pain n = 50) who were re-assessed 7 days after completing the initial survey.

**Copyright and licensing.** © 2002 Michael JL Sullivan. The IEQ-SF is licensed and distributed by Mapi Research Trust on behalf of the copyright holder. The IEQ-SF-J was developed under license from the copyright holder. Permissions and conditions of use are available through ePROVIDE (https://eprovide.mapi-trust.org).

**Table S3. Test–retest reliability of the IEQ-SF-J under alternative definitions of clinical stability**

| Stability definition | n | ICC(3,1) | 95% CI |
|---|---|---|---|
| All paired respondents | 100 | 0.60 | 0.46–0.71 |
| Stable pain intensity: absolute NRS change ≤1 | 61 | 0.66 | 0.49–0.78 |
| Stable pain intensity and disability: absolute NRS change ≤1 and PDI-5 mean item-score change ≤1 | 31 | 0.68 | 0.43–0.83 |

**Abbreviations.** CI, confidence interval; ICC, intraclass correlation coefficient; NRS, Numeric Rating Scale; PDI-5, 5-item Pain Disability Index.
**Note.** Clinical stability in pain intensity was defined as an absolute change of no more than 1 point on the 0–10 NRS. Stability in pain-related disability was defined as an absolute change of no more than 1 point in the PDI-5 mean item score (equivalent to no more than 5 points on the 0–50 summed score). A stricter criterion requiring no change in both measures identified only three participants and was therefore not used for reliability estimation.

**Table S4. Spearman correlation coefficients and 95% confidence intervals**

| | 1 | 2 | 3 | 4 | 5 | 6 | 7 | 8 |
|---|---|---|---|---|---|---|---|---|
| **Headache** | | | | | | | | |
| **1. IEQ-SF** | - | | | | | | | |
| **2. IEQ** | 0.60 [0.52 to 0.66] | - | | | | | | |
| **3. PCS** | 0.63 [0.55 to 0.69] | 0.68 [0.61 to 0.73] | - | | | | | |
| **4. PHQ-2** | 0.32 [0.21 to 0.42] | 0.49 [0.40 to 0.57] | 0.37 [0.26 to 0.46] | - | | | | |
| **5. Demand** | 0.26 [0.15 to 0.36] | 0.26 [0.15 to 0.37] | 0.24 [0.13 to 0.34] | 0.23 [0.12 to 0.34] | - | | | |
| **6. Low control** | 0.17 [0.06 to 0.28] | 0.25 [0.14 to 0.35] | 0.22 [0.11 to 0.32] | 0.22 [0.11 to 0.33] | 0.25 [0.14 to 0.36] | - | | |
| **7. NRS** | 0.37 [0.27 to 0.46] | 0.33 [0.23 to 0.43] | 0.33 [0.23 to 0.43] | 0.23 [0.11 to 0.33] | 0.14 [0.02 to 0.25] | 0.16 [0.04 to 0.27] | - | |
| **8. PDI** | 0.45 [0.35 to 0.53] | 0.43 [0.34 to 0.52] | 0.44 [0.35 to 0.53] | 0.25 [0.15 to 0.36] | 0.17 [0.05 to 0.27] | 0.18 [0.07 to 0.29] | 0.44 [0.34 to 0.52] | - |
| **9. AIS-5** | 0.36 [0.25 to 0.45] | 0.39 [0.29 to 0.48] | 0.34 [0.23 to 0.43] | 0.45 [0.35 to 0.53] | 0.23 [0.12 to 0.34] | 0.25 [0.14 to 0.35] | 0.33 [0.22 to 0.42] | 0.31 [0.20 to 0.41] |

| | 1 | 2 | 3 | 4 | 5 | 6 | 7 | 8 |
|---|---|---|---|---|---|---|---|---|
| **Low back pain** | | | | | | | | |
| **1. IEQ-SF** | - | | | | | | | |
| **2. IEQ** | 0.67 [0.60 to 0.73] | - | | | | | | |
| **3. PCS** | 0.71 [0.65 to 0.77] | 0.74 [0.69 to 0.79] | - | | | | | |
| **4. PHQ** | 0.38 [0.28 to 0.48] | 0.51 [0.41 to 0.59] | 0.41 [0.31 to 0.50] | - | | | | |
| **5. Demand** | 0.20 [0.09 to 0.31] | 0.12 [0.01 to 0.23] | 0.16 [0.04 to 0.27] | 0.03 [-0.08 to 0.15] | - | | | |
| **6. Low control** | 0.17 [0.06 to 0.28] | 0.26 [0.15 to 0.36] | 0.16 [0.05 to 0.27] | 0.19 [0.08 to 0.30] | 0.30 [0.19 to 0.40] | - | | |
| **7. NRS** | 0.33 [0.22 to 0.42] | 0.31 [0.21 to 0.41] | 0.32 [0.22 to 0.42] | 0.18 [0.07 to 0.29] | 0.09 [-0.02 to 0.20] | 0.02 [-0.10 to 0.13] | - | |
| **8. PDI** | 0.55 [0.46 to 0.62] | 0.54 [0.45 to 0.61] | 0.58 [0.50 to 0.65] | 0.30 [0.20 to 0.40] | 0.09 [-0.03 to 0.20] | 0.13 [0.02 to 0.24] | 0.38 [0.28 to 0.47] | - |
| **9. AIS-5** | 0.31 [0.20 to 0.41] | 0.42 [0.32 to 0.51] | 0.45 [0.35 to 0.53] | 0.40 [0.30 to 0.49] | 0.13 [0.02 to 0.24] | 0.09 [-0.02 to 0.21] | 0.26 [0.15 to 0.36] | 0.36 [0.25 to 0.45] |

**Abbreviations.** AIS-5, Athens Insomnia Scale 5-item version; demand, job demands; IEQ, Injustice Experience Questionnaire; IEQ-SF, Injustice Experience Questionnaire-Short Form; low control, low job control (reversed); NRS, Numeric Rating Scale; PCS, Pain Catastrophizing Scale; PDI, Pain Disability Index; PHQ, Patient Health Questionnaire-2.

**Note.** Values are presented as Spearman's rank correlation coefficients [95% Confidence Interval]. All confidence intervals are 95%.

**Table S5. Item-level descriptive statistics, IRT parameters, and differential item functioning of the IEQ-SF-J**
**Panel A. Item-level descriptive statistics and IRT parameters**

| Item | Group | Mean | SD | Response Category<br>0 (never)<br>n (%) | Response Category<br>1 (sometimes)<br>n (%) | Response Category<br>2 (often)<br>n (%) | a | b1 | b2 | Floor Effect | Ceiling Effect | Factor loading (EFA) |
|---|---|---|---|---|---|---|---|---|---|---|---|---|
| **Item 1** | **Total** | 1.05 | 0.56 | 82 (13.7) | 408 (68.0) | 110 (18.3) | 2.05 | -1.44 | 1.17 | No | Yes | 0.622 |
| | **Headache** | 1.07 | 0.60 | 45 (15.0) | 190 (63.3) | 65 (21.7) | 2.25 | -1.30 | 0.98 | Yes | Yes | 0.674 |
| | **Low back pain** | 1.03 | 0.52 | 37 (12.3) | 218 (72.7) | 45 (15.0) | 2.06 | -1.52 | 1.36 | No | Yes | 0.579 |
| **Item 2** | **Total** | 0.73 | 0.62 | 218 (36.3) | 325 (54.2) | 57 (9.5) | 2.14 | -0.44 | 1.68 | Yes | No | 0.670 |
| | **Headache** | 0.89 | 0.64 | 80 (26.7) | 173 (57.7) | 47 (15.7) | 2.20 | -0.79 | 1.27 | Yes | Yes | 0.688 |
| | **Low back pain** | 0.57 | 0.56 | 138 (46.0) | 152 (50.7) | 10 (3.3) | 1.91 | -0.12 | 2.50 | Yes | No | 0.606 |
| **Item 3** | **Total** | 0.63 | 0.67 | 286 (47.7) | 248 (41.3) | 66 (11.0) | 1.82 | -0.06 | 1.70 | Yes | No | 0.641 |
| | **Headache** | 0.76 | 0.67 | 112 (37.3) | 148 (49.3) | 40 (13.3) | 1.70 | -0.44 | 1.59 | Yes | No | 0.615 |
| | **Low back pain** | 0.51 | 0.65 | 174 (58.0) | 100 (33.3) | 26 (8.7) | 1.87 | 0.29 | 1.87 | Yes | No | 0.643 |
| **Item 4** | **Total** | 0.68 | 0.65 | 255 (42.5) | 285 (47.5) | 60 (10.0) | 2.47 | -0.22 | 1.57 | Yes | No | 0.730 |
| | **Headache** | 0.77 | 0.67 | 110 (36.7) | 150 (50.0) | 40 (13.3) | 2.13 | -0.43 | 1.43 | Yes | No | 0.695 |
| | **Low back pain** | 0.58 | 0.61 | 145 (48.3) | 135 (45.0) | 20 (6.7) | 2.99 | -0.03 | 1.73 | Yes | No | 0.767 |
| **Item 5** | **Total** | 0.51 | 0.63 | 340 (56.7) | 216 (36.0) | 44 (7.3) | 4.62 | 0.19 | 1.54 | Yes | No | 0.818 |
| | **Headache** | 0.61 | 0.67 | 150 (50.0) | 118 (39.3) | 32 (10.7) | 4.72 | 0.01 | 1.31 | Yes | No | 0.831 |
| | **Low back pain** | 0.41 | 0.57 | 190 (63.3) | 98 (32.7) | 12 (4.0) | 4.11 | 0.39 | 1.88 | Yes | No | 0.783 |

**Abbreviation.** SD = standard deviation; a, discrimination; b1, threshold parameter 1; b2, threshold parameter 2.
**Note.** The IEQ-SF-J comprises 5 items rated on a 3-point scale: 0 = never, 1 = sometimes, 2 = often. Floor or ceiling effects are considered present ("Yes") if ≥ 15% of the respondents selected the lowest or highest response category, respectively. The discrimination parameter (a) reflects how well an item differentiates between individuals with different levels of the underlying latent trait (perceived injustice). A higher value indicates better discrimination; values > 1.70 are typically considered "very high" discrimination. The threshold parameters ($b_1$ and $b_2$) represent the point on the latent trait continuum (in standard deviation units) where a respondent has a 50% probability of selecting the higher response category or above. Specifically, $b_1$ separates "0 (never)" from "1 (sometimes) and 2 (often)", while $b_2$ separates "0 (never) and 1 (sometimes)" from "2 (often)". Total N = 600, headache n = 300, low back pain n = 300.

**Panel B. Differential item functioning across headache and low back pain subgroups**

| Item | Uniform DIF, p | Non-uniform DIF, p | Overall DIF, BH-FDR q | ΔNagelkerke $R^2$ (overall) |
|---|---|---|---|---|
| 1 | 0.0042 | 0.1560 | 0.0102 | 0.0140 |
| 2 | <0.0001 | 0.2403 | <0.0001 | 0.0279 |
| 3 | 0.0140 | 0.0399 | 0.0102 | 0.0133 |
| 4 | 1.0000 | 0.0783 | 0.2654 | 0.0036 |
| 5 | 0.9203 | 0.3116 | 0.5963 | 0.0010 |

**Abbreviations.** BH-FDR, Benjamini–Hochberg false discovery rate; DIF, differential item functioning.
**Note.** Uniform DIF was assessed by adding pain subgroup to an ordinal logistic model conditioned on the four-item rest score; non-uniform DIF was assessed by adding the subgroup × rest-score interaction. Overall DIF q values were adjusted across the five items using the Benjamini–Hochberg procedure. ΔNagelkerke $R^2$ represents the total item-level DIF effect size. Items 1–3 showed statistically detectable DIF after FDR correction, but ΔNagelkerke $R^2$ values were small for all five items (0.0010–0.0279).

**Table S6. Full SEM results for direct, indirect, and total associations of job demands and low job control with pain interference, depressive symptoms, and insomnia**

| Outcome | Sample | Predictor | Fit index (RMSEA/CFI) | Direct effect on outcome | Indirect effect involving IEQ-SF, β (95% BC bootstrap CI) | Total effect on outcome | Interpretation |
|---|---|---|---|---|---|---|---|
| **PDI** | Overall | Job demand | 0.065 / 0.980 | B = -0.241, p = 0.338 | 0.133 (0.073 to 0.192) | — | Indirect-only association pattern |
| | | Low job control | | B = 0.241, p = 0.322 | 0.082 (0.028 to 0.142) | — | Indirect-only mediation |
| **PDI** | Headache | Job demand | 0.073 / 0.974 | B = 0.241, p = 0.705 | 0.134 (0.041 to 0.227)† | B = 1.216, p = 0.014 | Indirect-only pattern; interpret cautiously† |
| | | Low job control | | B = -0.721, p = 0.229 | 0.131 (0.043 to 0.312)† | B = 0.163, p = 0.728 | Weak/inconclusive indirect association pattern; interpret cautiously† |
| **PDI** | Low back pain | Job demand | 0.056 / 0.982 | B = -0.372, p = 0.250 | 0.111 (0.021 to 0.205) | B = 0.296, p = 0.429 | Indirect-only mediation |
| | | Low job control | | B = 0.118, p = 0.695 | 0.098 (0.014 to 0.182) | B = 0.670, p = 0.048 | Indirect-only mediation; significant total effect |
| **PHQ-2** | Overall | Job demand | 0.053 / 0.985 | B = 0.006, p = 0.722 | 0.084 (0.046 to 0.130) | B = 0.039, p = 0.022 | Indirect-only mediation |
| | | Low job control | | B = 0.048, p = 0.003 | 0.052 (0.017 to 0.092) | B = 0.067, p < 0.001 | Direct and indirect association pattern |
| **PHQ-2** | Headache | Job demand | 0.050 / 0.987 | B = 0.039, p = 0.106 | 0.071 (0.027 to 0.132) | B = 0.068, p = 0.005 | Indirect-only mediation |
| | | Low job control | | B = 0.060, p = 0.010 | 0.037 (0.002 to 0.092) | B = 0.074, p = 0.001 | Predominantly direct association |

| | | | | | | | |
|---|---|---|---|---|---|---|---|
| **PHQ-2** | Low back pain | Job demand | 0.053 / 0.983 | B = -0.031, p = 0.163 | 0.068 (0.014 to 0.133) | B = -0.007, p = 0.766 | Indirect-only mediation |
| | | Low job control | | B = 0.038, p = 0.061 | 0.060 (0.010 to 0.119) | B = 0.058, p = 0.006 | Indirect-only mediation |
| **AIS-5** | Overall | Job demand | 0.055 / 0.985 | β = 0.066, p = 0.126 | 0.089 (0.049 to 0.135) | B = 0.219, p = 0.001 | Indirect-only mediation |
| | | Low job control | | β = 0.116, p = 0.011 | 0.055 (0.018 to 0.100) | B = 0.225, p < 0.001 | Direct and indirect association pattern |
| **AIS-5** | Headache | Job demand | 0.043 / 0.991 | B = 0.086, p = 0.314 | 0.092 (0.040 to 0.158) | — | Indirect-only mediation |
| | | Low job control | | B = 0.256, p = 0.003 | 0.048 (marginal; non-standardized indirect effect: B = 0.065, 95% BC bootstrap CI: 0.003 to 0.157, p = 0.089)‡ | B = 0.321, p < 0.001 | Predominantly direct association; indirect effect marginal |
| **AIS-5** | Low back pain | Job demand | 0.057 / 0.980 | B = 0.081, p = 0.330 | 0.062 (0.017 to 0.170) | B = 0.162, p = 0.071 | Indirect-only mediation |
| | | Low job control | | B = 0.056, p = 0.482 | 0.055 (0.011 to 0.142) | B = 0.122, p = 0.128 | Indirect-only mediation |

**Abbreviations:** AIS-5, Athens Insomnia Scale 5-item version; BC, bias-corrected; IEQ-SF, short-form Injustice Experience Questionnaire; PHQ-2, Patient Health Questionnaire-2; PDI, Pain Disability Index; SEM, structural equation modeling.

**Note.** Indirect effects are shown as standardized coefficients (β) with 95% bias-corrected bootstrap confidence intervals based on 5,000 resamples, except where otherwise noted. Direct and total effects are shown using the coefficients explicitly available from the model output excerpts. In several models, the pasted excerpts did not retain standardized STDYX values for direct and total effects; therefore, the reported coefficients for those cells are presented as available rather than forcibly standardized. Higher values of low job control indicate lower perceived job control. For the PDI model in the headache subgroup, bootstrap estimation was unstable across repeated runs; therefore, the indirect effects in this subgroup should be interpreted with caution. For the AIS model in the headache subgroup, the indirect effect of low job control via latent IEQ-SF was marginal (standardized indirect effect β = 0.048), while the corresponding non-standardized indirect effect was B = 0.065 (95% BC bootstrap CI: 0.003 to 0.157, p = 0.089).

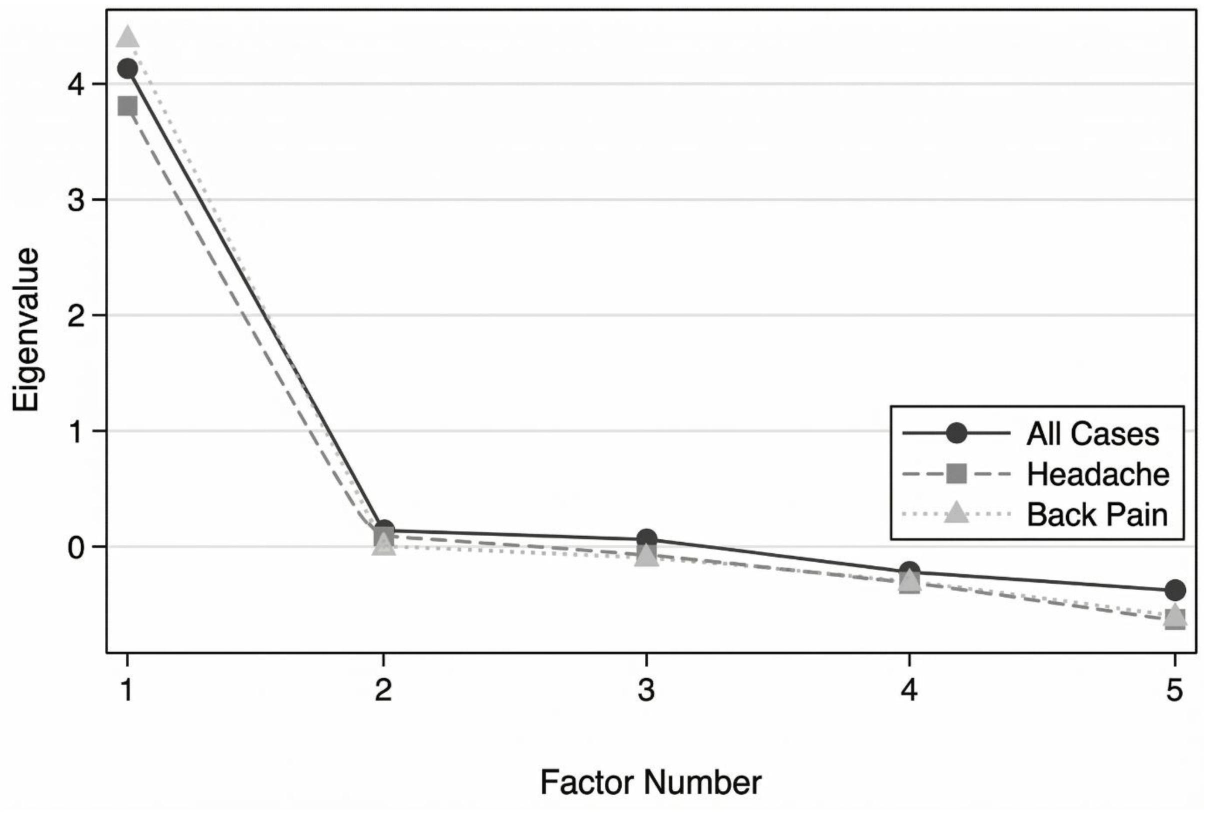


**Figure S1. Scree plot of eigenvalues across varying numbers of factors for the total sample (All Cases), headache subgroup, and low back pain subgroup.** The sharp drop following the first factor visually supports the unidimensional structure of the IEQ-SF-J across all conditions.

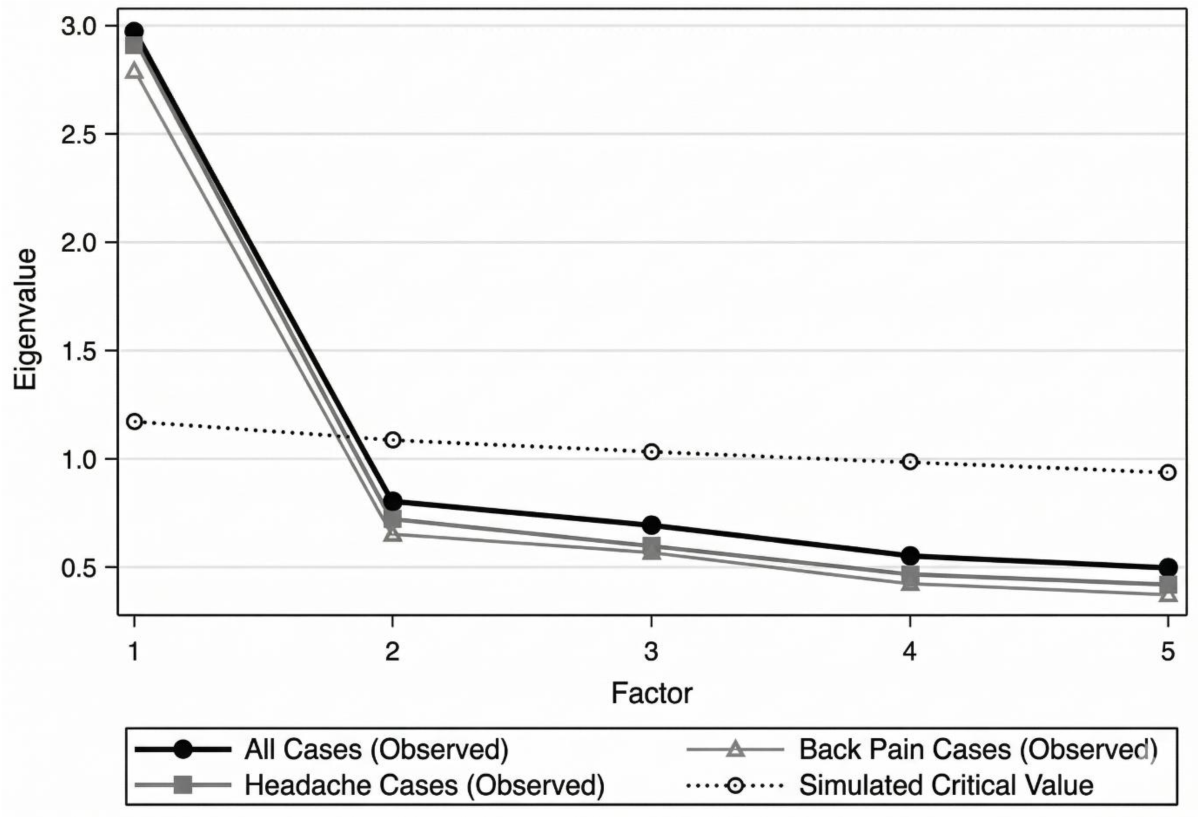


**Figure S2. Parallel analysis comparing observed eigenvalues against simulated critical values for the total sample (All Cases), headache subgroup, and low back pain subgroup.** Only the first factor's observed eigenvalue exceeds the simulated critical value, confirming the retention of a single latent factor for the IEQ-SF-J.
